# Rethinking antiviral effects for COVID-19 in clinical studies: early initiation is key to successful treatment

**DOI:** 10.1101/2020.05.30.20118067

**Authors:** Shoya Iwanami, Keisuke Ejima, Kwang Su Kim, Koji Noshita, Yasuhisa Fujita, Taiga Miyazaki, Shigeru Kohno, Yoshitsugu Miyazaki, Shimpei Morimoto, Shinji Nakaoka, Yoshiki Koizumi, Yusuke Asai, Kazuyuki Aihara, Koichi Watashi, Robin N. Thompson, Kenji Shibuya, Katsuhito Fujiu, Alan S. Perelson, Shingo Iwami, Takaji Wakita

## Abstract

Development of an effective antiviral drug for COVID-19 is a global health priority. Although several candidate drugs have been identified through *in vitro* and *in vivo* models, consistent and compelling evidence for effective drugs from clinical studies is limited. The lack of evidence could be in part due to heterogeneity of virus dynamics among patients and late initiation of treatment. We first quantified the heterogeneity of viral dynamics which could be a confounder in compassionate use programs. Second, we demonstrated that an antiviral drug is unlikely to be effective if initiated after a short period following symptom onset. For accurate evaluation of the efficacy of an antiviral drug for COVID-19, antiviral treatment should be initiated before or soon after symptom onset in randomized clinical trials.

**One Sentence Summary:** Study design to evaluate antiviral effect.

---

Development of an effective antiviral drug for COVID-19 is a global health priority. Along with the development of new antiviral drugs, repurposing existing drugs for COVID-19 treatment is accelerated (*1*). Some antiviral drugs have shown high efficacy against Severe Acute Respiratory Syndrome Coronavirus 2 (SARS-CoV-2) both *in vitro* and *in vivo* models (*2, 3*). A number of clinical studies such as compassionate use programs and clinical trials have been conducted or are underway to test the efficacy of FDA-approved drugs, such as lopinavir and ritonavir, chloroquine, favipiravir, and remdesivir (RDV) (*4–7*). However, the results from those clinical studies were often non-significant and sometimes inconsistent. This may be in part attributable to non-rigorous study design, which masks the true efficacy of antivirals (*8*). Clinical trial design usually takes months to formulate the study protocol (i.e., dose of drugs, clinical outcomes to be evaluated, sample size, assessment of safety), and requires collecting preliminary data. However, the urgent need to find effective antiviral treatment for COVID-19 may have led to rushed studies.

In compassionate use programs (i.e., observational studies), whether and when antiviral treatment is initiated is determined by health practitioners along with patients and their next of kin, therefore, there are potential confounders, such as the patients’ clinical characteristics and pre-existing conditions that influence both treatment-control allocation and clinical outcomes. As a consequence, conclusions from the program could be biased even when all observable confounders are addressed in the analysis (*9*). However, the programs are widely used for hypothesis building. Contrary to compassionate use programs, clinical trials, particularly randomized clinical trials, are considered robust against confounder effects and the most reliable study design, although designing such trials requires several time-consuming steps (developing and registration of the protocol). **Table S1** summarizes the current major clinical studies for antiviral treatment of SARS-CoV-2. Indeed, the results from these clinical studies have yielded null or inconsistent findings.

To help understand the mechanism behind this, we introduce a mathematical model to describe the within-host viral dynamics of SARS-CoV-2 and illustrate its usefulness by analyzing viral load data from clinical studies employing our developed quantitative approaches (*10–12*). Here, we demonstrate that at least two factors can mask the effect of antivirals in clinical studies for COVID-19: 1) heterogeneity of virus dynamics among patients and 2) late timing of treatment initiation. We also propose a novel approach to calculate sample size (i.e., the required or minimum sample size to infer whether or not the antiviral is effective assuming the drug is truly effective) accounting for within host virus dynamics.

SARS-CoV-2 viral load data were analyzed using a mathematical model (see **Supplementary Text**) to quantify the heterogeneity in viral dynamics among patients and examine its source. Longitudinal viral load data from 38 patients from different countries were fitted simultaneously using a nonlinear mixed-effect modelling approach (see **Materials and Methods**). With the estimated parameters for each patient (listed in **Table S2**), viral load since symptom onset were fully reconstructed even when viral load was not measured or under the detection limit (see **Fig.1A** and **Fig.S1**), which enabled quantitative comparison of viral load dynamics between patients. The reconstructed viral loads over time were analyzed with a clustering approach (see **Fig.1B** and **Materials and Methods**), and placed into three groups. To understand the source of difference in viral dynamics between groups, we tested for differences in the estimated parameters (i.e., *β*, *γ*, *δ* and *V*(0). See **Materials and Methods**) among groups. Statistically significant between-group differences were found in the initial viral load at symptom onset, *V*(0), and the death rate of virus producing cells per day, *δ* (**Fig.S2**). The difference in *δ* manifests itself in the speed of viral load decay, that is, a small value of *δ* corresponds to a slow decay in viral load (see **Fig.1B**). We named the three groups as rapid (*δ* > 0.75), medium (0.4 < *δ* < 0.75), and slow (*δ* < 0.4) viral load decay groups (see **Fig.2A**). If a patient possesses strong viral defenses including immune mediated, the virus producing cells are removed quickly, which corresponds to a shorter duration of virus production and rapid viral load decay. The duration of the virus detection in respiratory samples has been associated with disease severity (*13*).

**Fig. 1.**
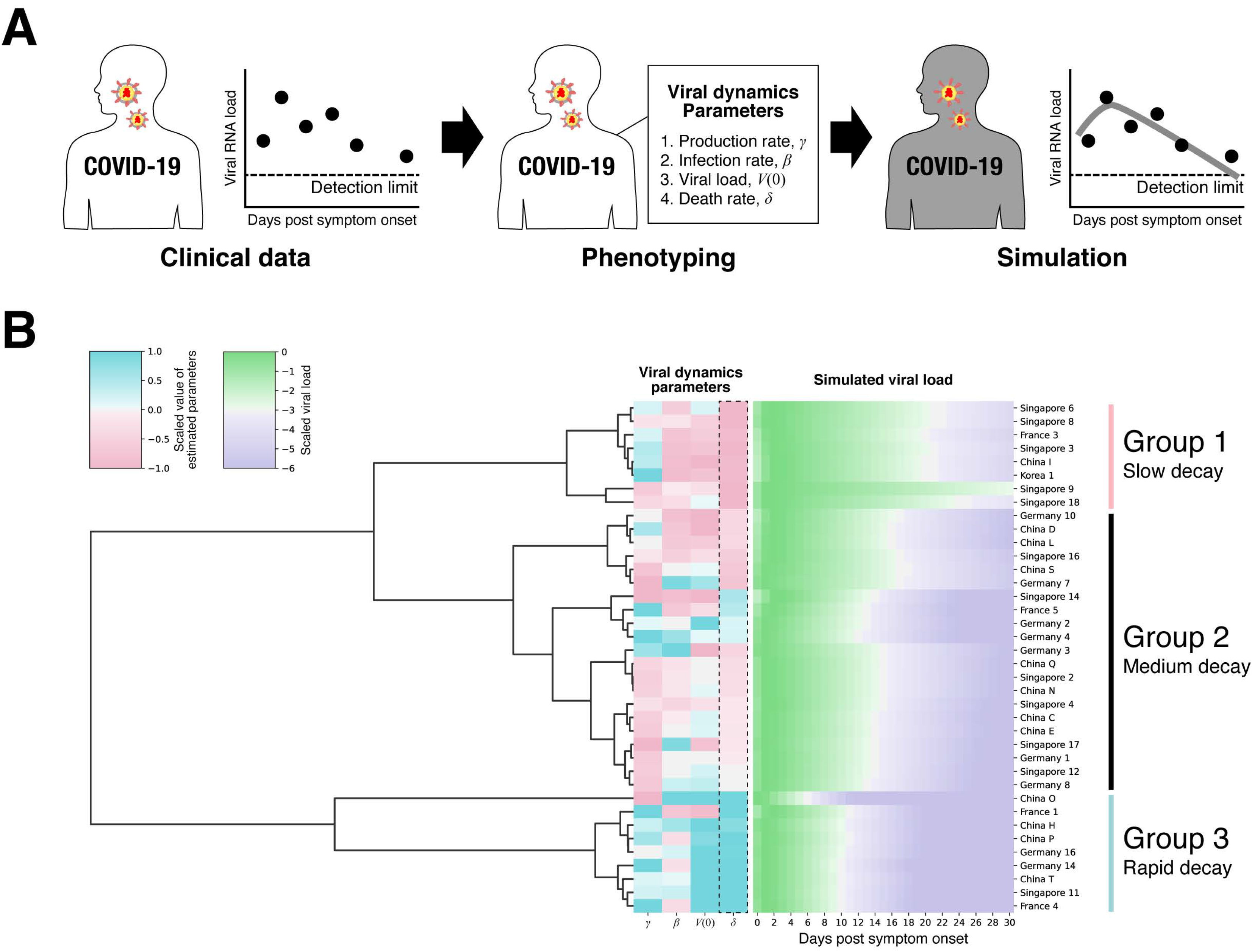
Phenotyping and clustering COVID-19 patients. (A) Datasets for measured SARS-CoV-2 RNA load of patients (i.e., clinical data) were analyzed by the mathematical model, and then virus dynamics parameters were estimated for each patient (i.e., phenotyping). Time-series data of viral load reconstructed based on estimated parameters were used to characterize patient variability. (B) Three different patient groups were identified by hierarchical clustering, and statistically significant between-groups differences were found in the the death rate of virus producing cell, *δ*.

**Fig. 2.**
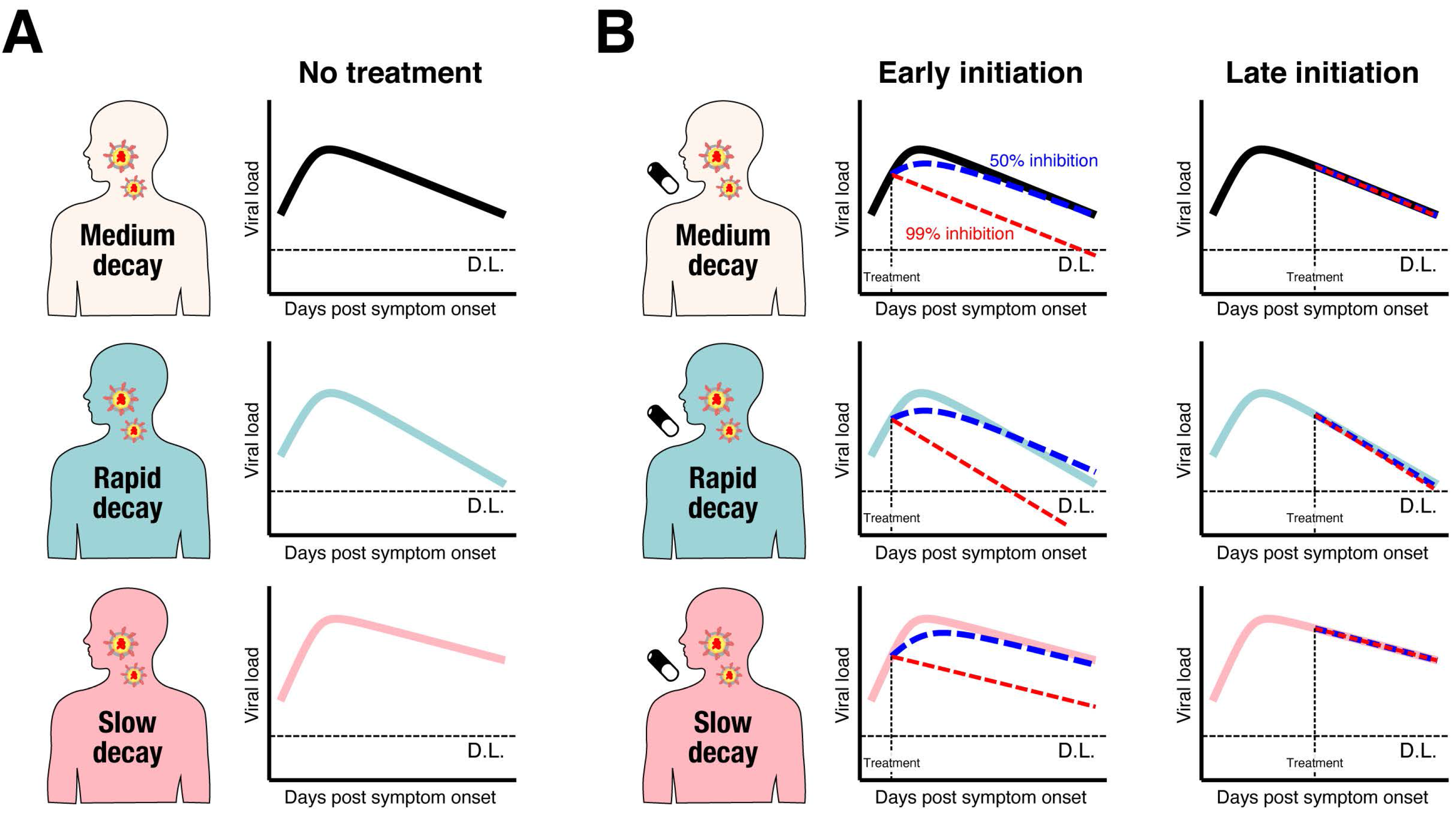
Patient variability and difference in therapeutic response. (A) Schematic representation of identified three different types of patients characterized by rapid, medium and slow viral load decay were shown. (B) Early initiation of antiviral drug which inhibits virus replication can change viral load decay depending on its inhibition rates and patient types. However, late initiation cannot influence the virus dynamics.

Based on our mathematical model and the estimated parameter distribution for each patient (**Table S2**), we conducted *in silico* experiments to determine the possible therapeutic response measured in terms of virus dynamics to investigate the expected outcome under drug treatments blocking virus replication (see **Supplementary Text**). Clinical outcomes are known to be related to the timing of antiviral treatment initiation in general and especially for influenza (*14–18*), and the antiviral effects are dependent on dose and the patients’ immune system (*19*). Thus, we studied several different scenarios varying the time of treatment initiation (0.5 or 5 days from symptom onset, which are before and generally after the estimated peak viral load in our dataset), inhibition rate (99% or 50%), and the three groups we have identified (rapid, medium, slow viral load decay). We found that early initiation of antiviral treatments with both high and low inhibition rates influence viral load decay, that is, viral load declines faster with high antiviral effect (see **Fig.2B** and **Fig.S3**). In contrast, the virus dynamics is not influenced if the treatment is initiated after the peak regardless of the inhibition rates and the patient types (see **Fig.2B** and **Fig.S3**). Note that these finding is not unique in SARS-CoV-2, but has been suggested in the other infectious diseases (*11, 20*).

**Fig. 3.**
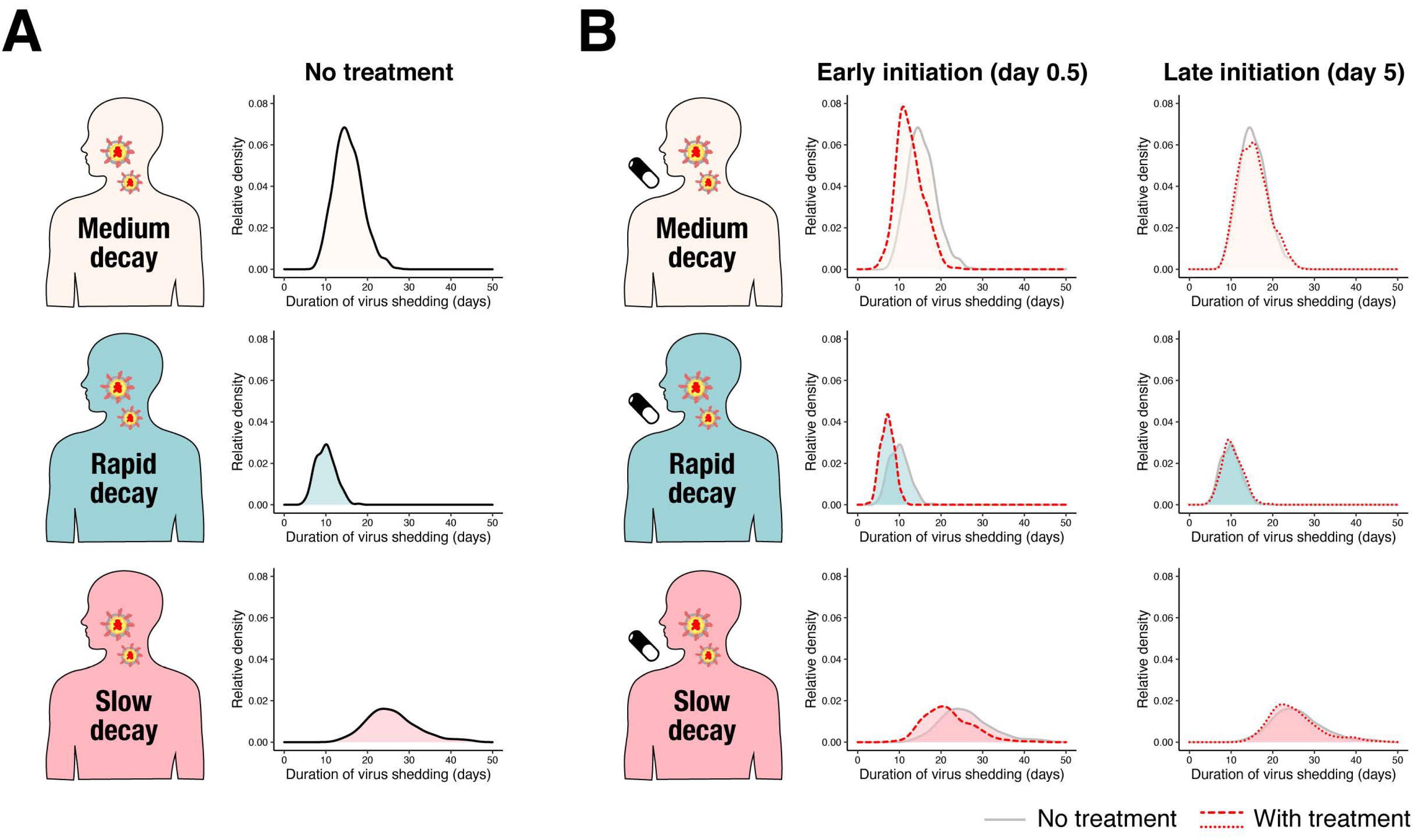
Duration of virus shedding in different types of patients. (A) Distributions of duration of virus shedding for the three groups (A) without treatment, and (B) with (early [Day 0.5] and late [Day 5] initiation of) treatment.

We further used the duration of virus shedding as an outcome, because it is one of the most frequently used outcomes to assess antiviral treatment for SARS-CoV-2 (see **Table S1**) (*4, 5, 7, 20*). The distribution of the duration under various conditions are shown in **Fig.3AB**. As we can expect from the difference in viral load dynamics, the duration is longer in the group with slow viral load decay (the averages in the groups with medium, rapid and slow decay were 15.3, 9.95 and 26.2 days, respectively). We further found early initiation of treatment significantly reduced the duration for all the groups, whereas late initiation did not change the duration (**Fig.3B and S4A**). As a sensitivity analysis we also computed and compared the cumulative viral load (area under the curve; AUC), which is another commonly used clinical outcome. We confirmed same trend on AUC as the duration (see **Supplementary Text, Fig.S4B** and **Fig.S5AB**).

**Fig. 4.**
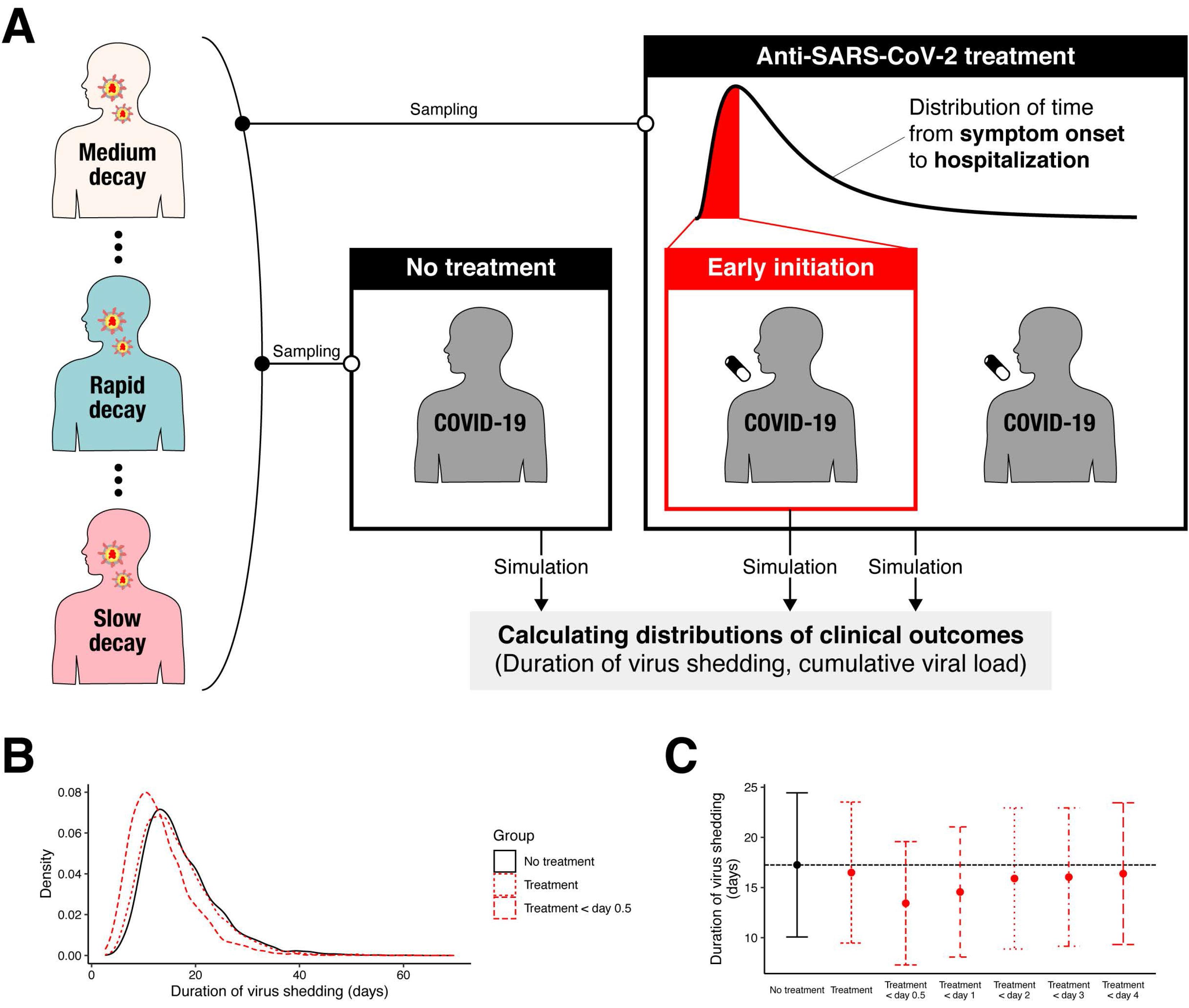
*In silico* randomized clinical trial of anti-SARS-CoV-2 in COVID-19 patients. (A) Simulation parameter sets were sampled from the distribution of estimated parameters for each patient type. The parameter sets were randomized into control or treatment groups. The treatment is initiated randomly following the distribution. Then the duration of virus shedding (and the cumulative viral load) were determined for each patient. (B) The distributions of duration of virus shedding for patients without and with treatment were shown in black and red curves, respectively. It was assumed that all patients (dotted) and the patients treated in 0.5 days from the onset of symptoms (dashed) were enrolled in *in silico* randomized clinical trials. (C) Means and standard deviation of the duration of virus shedding under different inclusion criteria were shown.

Compassionate use programs may not provide valid inference because it is not possible to control all confounders. For example, heterogeneous immune responses, partially represented by death rate of infected cells in the model, can confound the inference, however, quantifying the immune response is difficult and other confounding variables may still exist. On the other hand, randomized clinical trials may not be influenced by confounding variables and could provide valid inference (see **Fig.4**). However, the clinical trial for COVID-19 should consider the timing of treatment initiation in the design, which clearly influence the sample size.

We computed the sample size needed for 80% power to detect at the 0.05 significance level a difference between treatment and control groups assuming patients are randomized and treated (with antiviral or placebo) immediately after hospitalization (see **Fig.4A**) (see **Supplementary Text** for the detail of sample size calculation). The computed sample size (**Table S3**) was much larger than the sample size in the randomized clinical trials of antivirals for SARS-CoV-2 (see **Table S1**) if patients are enrolled regardless of the time of treatment initiation (**Fig.4BC**). This large sample size is needed given that the treatment is initiated 4.6 days after onset of symptoms on average, which is after the viral load peak. Next, we set an inclusion criterion based on the time from symptom onset to treatment initiation (**Fig.4BC**) and computed the sample size (**Table S3**). The required sample size is smaller with more strict (shorter time) inclusion criteria. Without exclusion criteria, 2720 patients per group are necessary to see a treatment effect using the duration of virus shedding as an outcome. If we enroll only the patients treated within 0.5 days from the onset of symptoms, the sample size is reduced to 98. There is a similar trend when AUC is used as an outcome (see **Table S3** and **Fig.S5CD**).

Developing effective antiviral drugs is the key for reducing the burden of COVID-19 on health care and avoiding stringent non-pharmaceutical interventions such as lockdown. Although the several candidate antivirals have been identified, randomized clinical trials demonstrating their effectiveness in reducing viral shedding are limited, and compassionate use programs demonstrated inconsistent results with the same antivirals. For example, compassionate use of hydroxychloroquine was reported in at least two papers, and the finding were not consistent. Gautret et al. reported significant antiviral efficacy (*21*); whereas Geleris et al. could not replicate the result (*22*). Mehra et al. found hydroxychloroquine or chloroquine treatment is associated with negative clinical outcomes. We identified heterogeneity in viral load kinetics characterized by the death rate of infected cells, which might be a confounding variable in compassionate use programs (**Fig.2**). We also found the timing of treatment initiation is non-linearly associated with the outcomes (duration of virus detection and AUC) (**Fig.3** and **Fig.S5**). If the treatment is initiated after the viral load peak, the antiviral effect would not be significant (**Fig.2**). Indeed, treatment initiation in most of the randomized clinical trials are way after the onset of symptoms (or not reported) (**Table S1**). For example, a significant effect of lopinavir-ritonavir (*4*) and RDV (*5*) was not observed in randomized clinical trials, which could be because treatments were initiated too late: after 13 and 11 days since the onset of symptoms, respectively.

We further searched clinical trials investigating antiviral efficacy registered in ClinicalTrials.gov. As of 22 May, 2020, 176 clinical trials were identified with the searching terms “antiviral” and “COVID”. Among them, 46 studies did not investigate the efficacy of antiviral drugs (anti-inflammatory drugs effect were investigated, for example), and 20 studies did not directly investigate antiviral effect (such as vaccine studies, safety studies). Among the remained 110 studies investigated antiviral effect, including remdesivir, chloroquine, and lopinavir/ritonavir, only 17 studies (15%) explicitly stated the time from symptom onset in inclusion/exclusion criteria, and average time from onset to randomization was 7.2 days, which is too late to observe antiviral effect.

The strength and uniqueness of our approach is that we accounted for viral dynamics in assessment of antivirals effect and sample size calculation. As far as we know, sample size calculation is dependent on the outcome distributions in control and treatment groups, in general. However, in the assessment of antiviral effect for SARS-CoV-2, this approach may not work, because the treatment effect is non-linearly dependent on timing of treatment initiation, which should be soon after the onset of symptoms.

There are several limitations in our approach. First, our within-host viral dynamics may not fully reflect the detailed physiological processes of virus replication of SARS-CoV-2. For example, our mathematical model assumed all target cells are homogeneous (i.e., single-target cell compartment). The susceptibility of target cells for SARS-CoV-2 infection is, however, depended on expression levels of its receptor, angiotensin converting enzyme 2: ACE2 (*23*), and therefore the susceptibility might be heterogeneous (i.e., multi-target cell compartments) even in the same organ. However, the viral dynamics of our model and that of the multi-target cell compartments will not be different unless a large fraction of total target cell in modified models remain uninfected around peak viral load. In principle, if relevant data exist, this could be included in an extended version of our model. Second, possible immunomodulation induced by the treatments was not modelled. That is, if anti-SARS-CoV-2 drugs induce immunomodulation as bystander effects, late initiation of treatments might still have the potential to reduce viral load, which is not reflected in our model (*18*).

Along with vaccines, developing effective antivirals is urgently needed. At present, most of the randomized clinical trials have failed to identify effective antivirals against SARS-CoV-2. However, this might not be because the antivirals are not effective, but because of imperfect design of the clinical studies. The timing of treatment initiation and virus dynamics should be accounted for in the study design (i.e., sample size and inclusion-exclusion criteria) to identify effective antivirals.

## Data Availability

The data examined in our manuscript were extracted from the published studies of SARS-CoV-2. We do not have the right to publicize the data, but we have cited those papers in the manuscript.

## Author contributions

Conceptualization, K.E., S.N., K.W., AS.P., S.I. T.W.; investigation, S.I., K.E., KS.K., S.N., Y.K., Y.F., K.A., RN.T., S.I.; supervision, K.E. and S.I.; writing, original draft, K.E., K.A., RN.T., K.S., A.S.P, and S.I.; writing-review and editing, all authors; funding acquisition: K.W., K.A., and S.I.; **Competing interests:** Authors declare no competing interests.;

## Data and materials availability

All data is available in the main text or the supplementary materials.

## Supplementary Materials

### This PDF file includes

Materials and Methods Supplementary Text Figs. S1 to S5

Tables S1 to S3

### Materials and Methods

#### Study data

The data examined in our manuscript were extracted from the published studies of SARS-CoV-2: Young et al. (*1*), Zou et al. (*2*), Wölfel et al. (*3*), Lescure et al. (*4*) and Kim et al. (*5*). To extract the data from the images in those papers, we used the software datathief III (version 1.5, Bas Tummers, http://www.datathief.org). We excluded patients who received antiviral treatment and for whom data were measured on only a single day (because a single time point data is not enough to estimate parameter values), and assumed that viral load values under the detection limit were set as the detection limit. We converted cycle threshold (Ct) values to viral RNA copies number values, where these quantities are inversely proportional to each other (*6*).

#### The nonlinear mixed effect model

A nonlinear mixed-effects model was used to fit the viral dynamics model to the viral load data. The model accounted for both a fixed effect (constant across patients) and a random effect (different between patients) in each parameter. Although the viral load was measured only a few times for some patients, the nonlinear mixed effects model makes the parameter estimation possible for those patients by using the data from the others. Further, we used this approach to account for individual variability. If the data are pooled and analyzed together, the between-patients variability will be lost, and the estimation of parameter is heavily influenced by patients with more observations. Thus, the parameter for patient *i*, *ϑ*_*i*_ (= *ϑ* × *e*^*π*^*i*) is represented as a product of P (a fixed effect) and *π*_*i*_ (a random effect). *π*_*i*_ follows the normal distribution with mean 0 and standard deviation Ω. Fixed effects and random effects were estimated using the stochastic approximation expectation-maximization algorithm and empirical Bayes’ method, respectively. The modes of the conditional distributions of parameters and initial values for each patient were summarized in **Table S2**. Fitting was performed using MONOLIX 2019R2 (http://www.lixoft.com) (*7*).

#### Clustering

Daily viral load values after the onset of symptoms were estimated by the model fitting. The estimated viral load data of each patient were rescaled by their maximum values, and log-transformed. Then, hierarchical clustering was performed on the data using the linkage function with Ward’s method () in SciPy (). We identified four major clusters of the viral dynamics after symptom onset. However, one group included only one patient, thus we decided to use three groups for further analyses by reassigning the patient to a closest group (**Fig. 1B**).

#### Statistical analysis

Estimated parameter distributions between the three groups with different viral load dynamics (i.e., rapid, medium, and slow viral load decay groups) were compared by ANOVA. Pairwise comparison was subsequently performed using Student’s t test. The p-values of the pairwise Student’s t test were adjusted by the Bonferroni correction.

### Supplementary Text

#### Mathematical model

COVID-19 dissemination among susceptible target cells is described by a mathematical model previously proposed by Kim et al. in (*8, 9*).

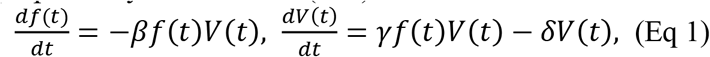

where *f*(*t*) is the relative fraction of uninfected target cells at time *t* to those at time 0 and *V*(*t*) is the amount of virus at time *t*, respectively. Both *f*(*t*) and *V*(*t*) are in linear scale. The parameters *β*, *γ*, and *δ* represent the rate constant for virus infection, the maximum rate constant for viral replication and the death rate of virus producing cells, respectively. All viral load data were simultaneously fitted using a nonlinear mixed-effect modelling approach, which estimates population parameters while accounting for inter-individual variation (see <u>The nonlinear mixed</u> <u>effect model</u>). The day from symptom onset was used as a time scale (i.e., *t* = 0 at symptom onset).

#### The outcomes characterize virus dynamics

We calculated the two quantities as outcomes: the duration of virus shedding from the onset of symptoms until the time the virus became undetectable (*T*_*D*_), and the cumulative viral load, i.e., the area under the curve of viral load 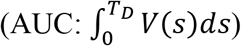. Both outcomes are expected to be reduced under effective antiviral treatment.

#### *In silico* experiments for anti-SARS-CoV-2 therapies

We investigated the antiviral effects of drugs blocking virus replication (such as lopinavir/ritonavir [HIV protease inhibitors], remdesivir [originally developed for hepatitis C, and considered potentially useful for Ebola virus], and the other nucleoside analogues (*10, 11*)) using the virus dynamics model with antiviral effect. The model under antiviral treatment initiated at *t*^∗^ days after symptom onset was developed based on the model (Eq 1) as follows:

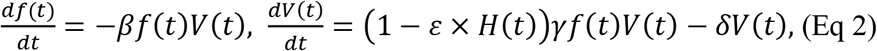

where *H*(*t*) is a Heaviside function indicating off- and on-treatment, defined as *H*(*t*) = 0 if *t* < *t*^∗^ (i.e., before treatment initiation): otherwise *H*(*t*) = 1. *ɛ* is the fraction of virus production inhibited by the therapy (0 < *ε* ≤ 1). *ε* = 1 when the virus replication from the infected cells are perfectly inhibited. We evaluated the expected antiviral effect of the treatment on the outcomes (duration of virus shedding and cumulative viral load) under different inhibition rates (*ε*) and initiation timings (*t*^∗^).

#### Sample size calculation

The allocation ratio is assumed as 1:1 (control:treatment). 5,000 parameter sets were randomly sampled from the estimated parameter distributions to create the treatment and the control groups (i.e., 10,000 parameter sets in total). For the treatment group, the treatment (99% inhibition rate on viral replication) was initiated following the distribution of time from onset to hospitalization, obtained from Bi et al.: *lognorm*(1.23, 0.79) (the mean is 4.64 [days]) (*12*), where the treatment was assumed to be initiated immediately after hospitalization. Thus, we obtained 5,000 outcomes for each group (duration of virus shedding in linear scale and cumulative viral load in log scale: **Fig.3BC** and **Fig.S5CD**, respectively). The sample size was computed using two-tailed Welch’s t test with significance level and power as 0.05 and 80%, respectively.

---

**Figure S1.**
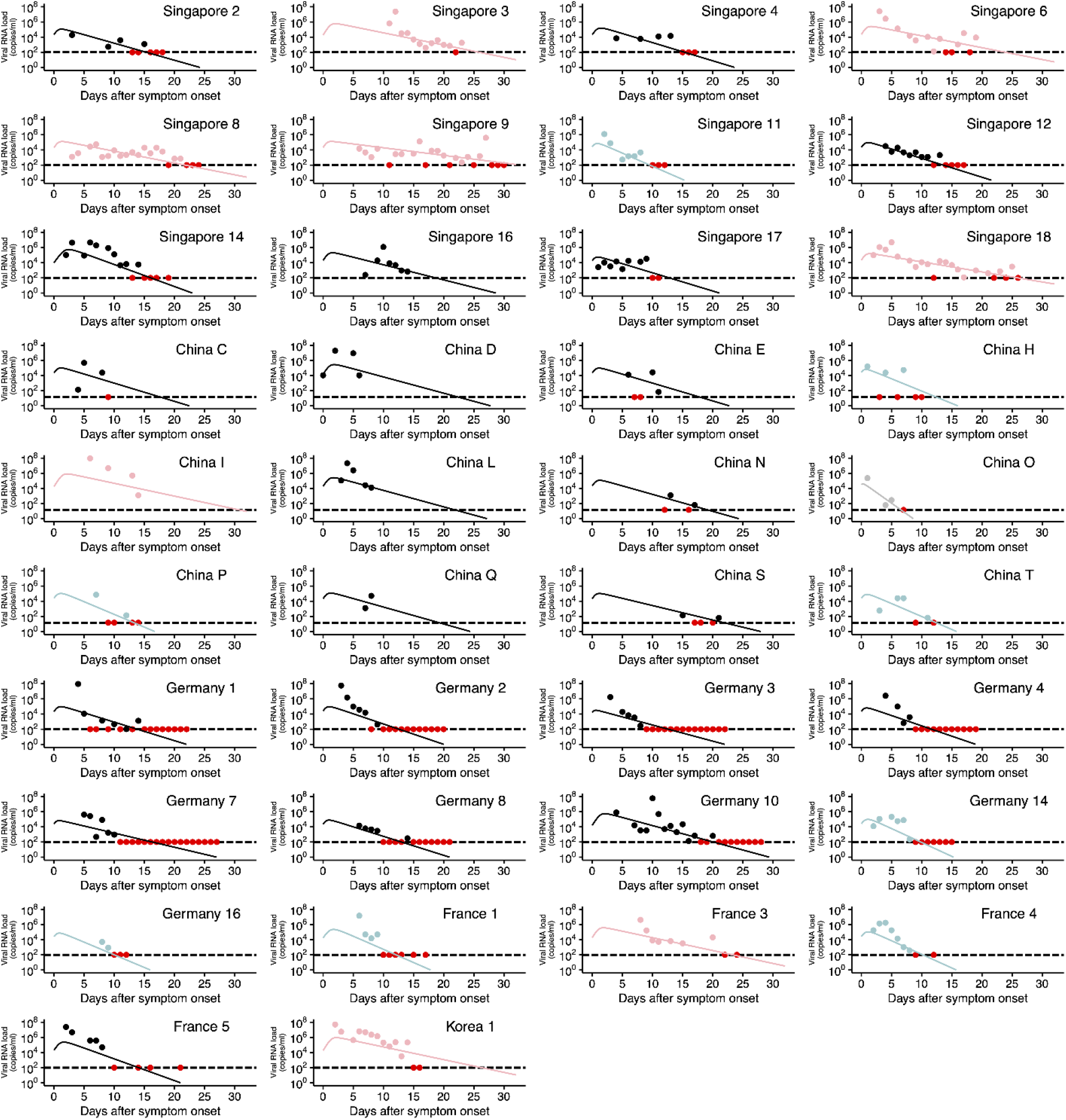
Observed and estimated viral load data for individual patients. Viral loads were measured using nasal swabs (China) and nasopharyngeal swabs (Singapore, Germany, France, and Korea) for SARS-CoV-2 infected patients since hospitalization. Note that the detection limits of the PCR assay for SARS-CoV-2 were 14.6 copies/ml (China) and 100 copies/ml (Singapore, Germany, France and Korea), respectively, and shown as dotted horizontal lines. The closed dots and curves correspond to the observed and the estimated viral load. Different colors of the dots and the lines (light blue, black, and pink) correspond to the three different types of patients characterized by rapid, medium and slow viral load decay, respectively. The red dots represent the data at or under the detection limit regardless of the group. Patient IDs are the same as in the original papers if available.

**Figure S2.**
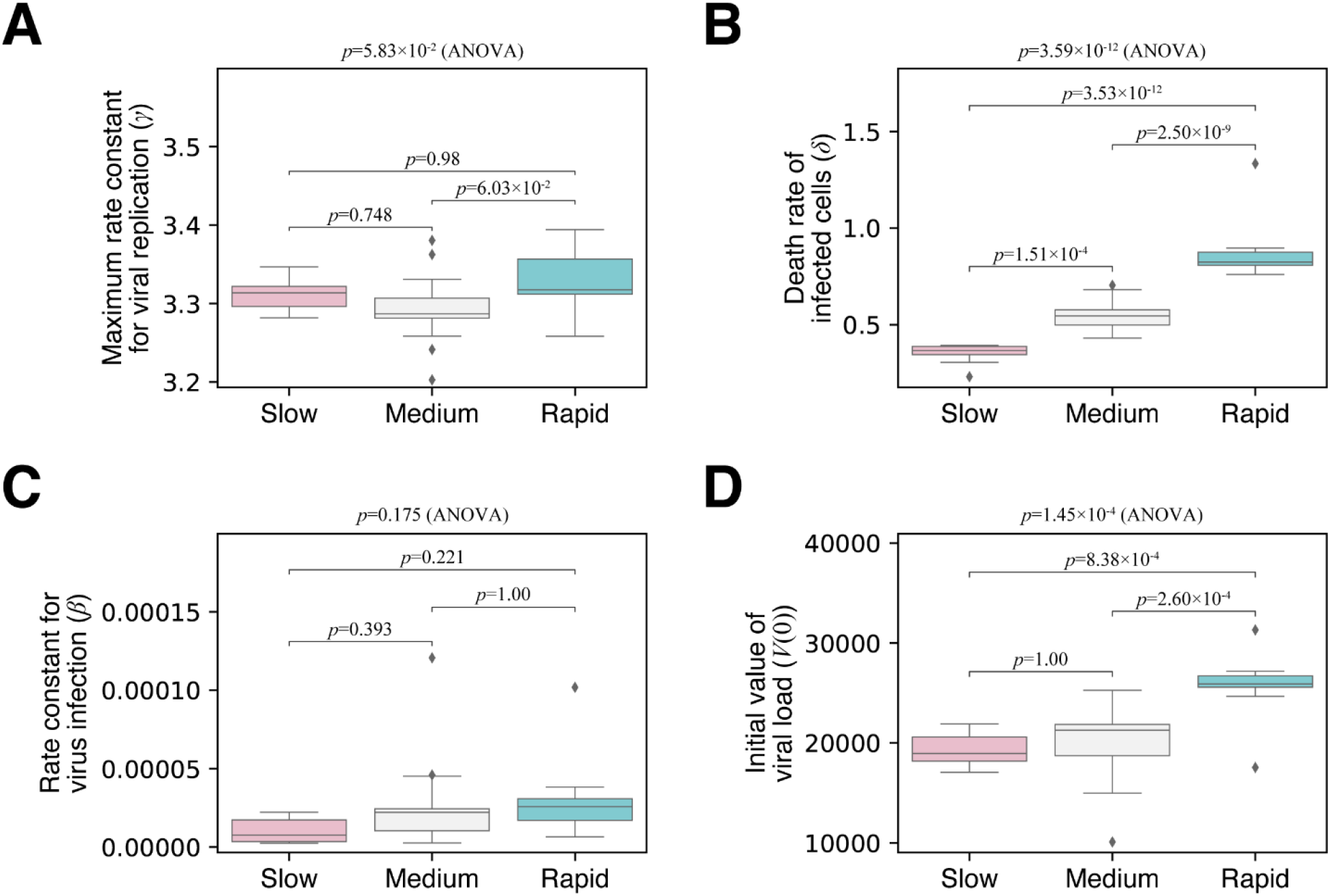
Estimated parameters of the mathematical model in the three different groups. Boxplots of estimates of (A) the rate constant for virus infection, *β*, (B) the maximum rate constant for viral replication, *γ*, (C) the death rate of virus producing cells, *δ*, and (D) viral load at symptom onset, *V*(0). Estimated parameter distributions between the three groups with different viral load dynamics (slow, medium, and rapid viral load decay groups) were compared by ANOVA. Pairwise comparison was subsequently performed using Student’s t test. The p-values of the pairwise Student’s t test were adjusted by the Bonferroni correction.

**Figure S3.**
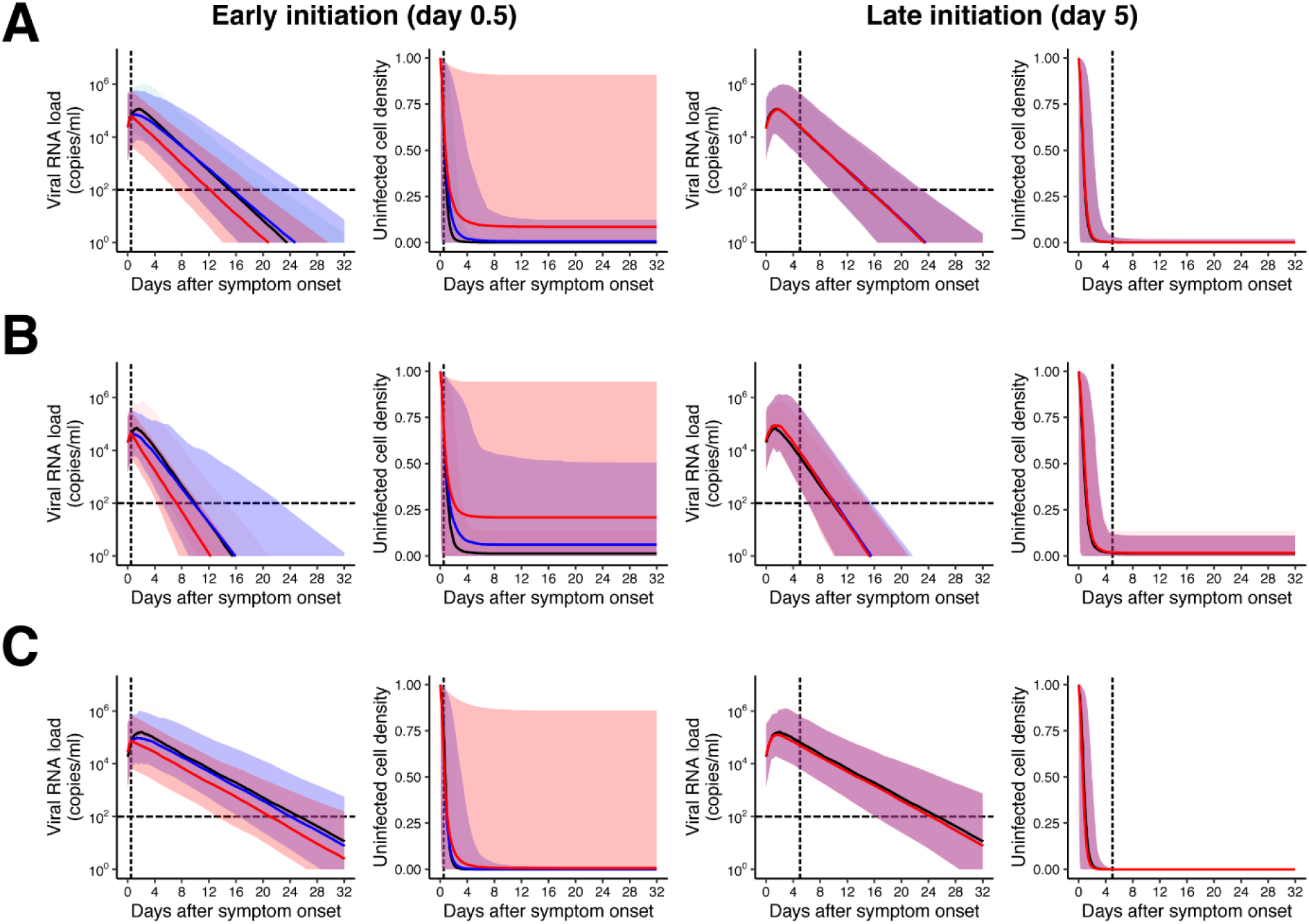
Expected virus dynamics under the antiviral treatment blocking viral replication. The antiviral treatment was assumed to be initiated after 0.5 or 5 days (named early and late initiations, respectively) from symptom onset with 99% and 50% inhibition rates (named high and low antiviral effects, respectively) for patients with (A) medium, (B) rapid, (C) slow viral load decay. Left and right panels in each group show the viral loads, *V*(*t*), and the relative fraction of uninfected target cells, *f*(*t*). The black and colored solid lines correspond to the mean of the values without and with the therapies (red: high, blue: low antiviral effects), respectively. The shadowed regions correspond to the 95% predictive intervals.

**Figure S4.**
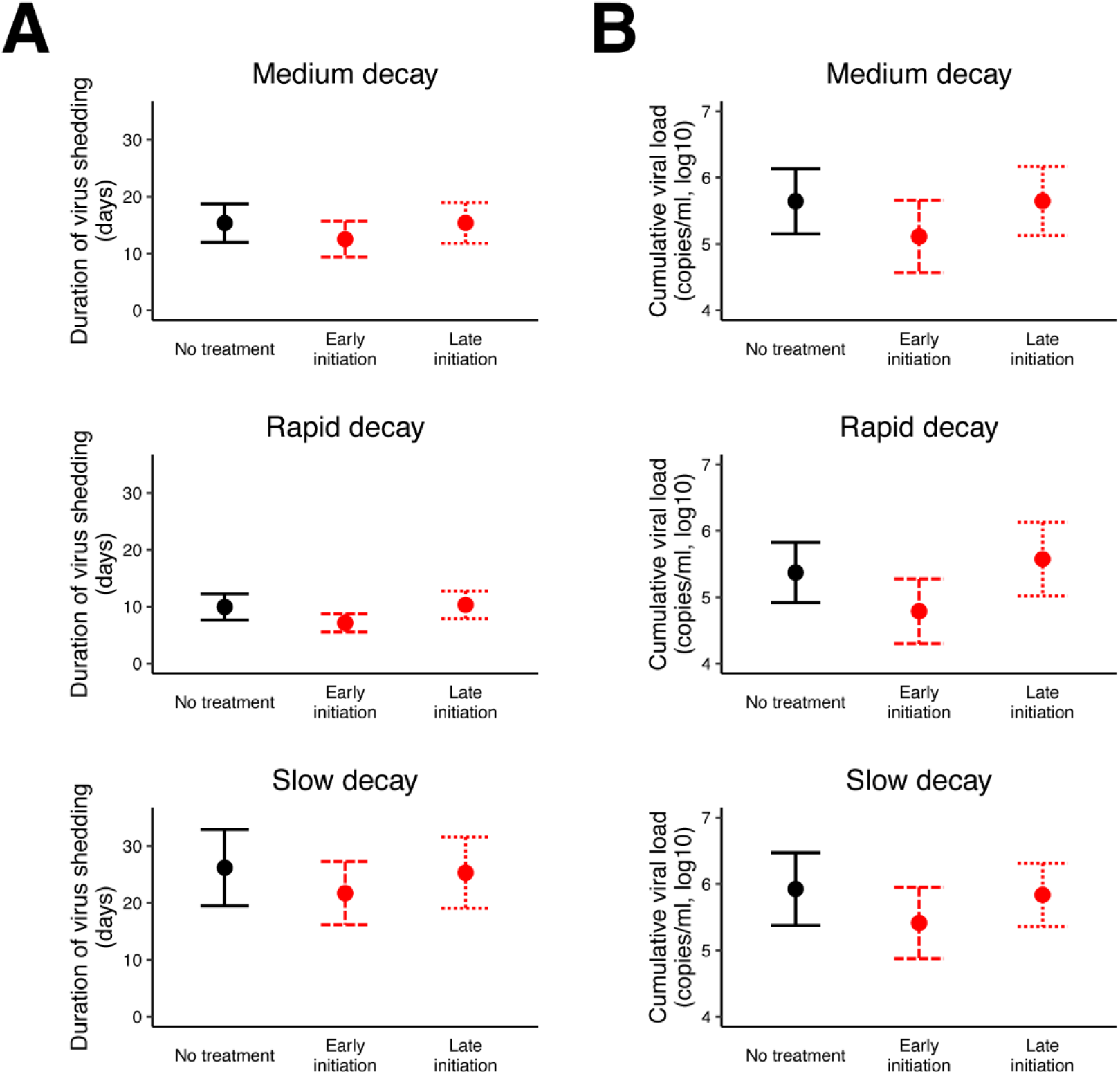
Distribution of duration of virus shedding and cumulative viral load. (A) Duration of virus shedding and (B) Cumulative viral load (in log scale) of SARS-CoV-2 for different types of patients (medium, rapid, and slow decay) with and without treatment. ‘Early initiation’ and ‘Late initiation’ means the early and the late treatment initiation (0.5 or 5 days after symptom onset). The dots and error bars represent the mean and the standard deviation.

**Figure S5.**
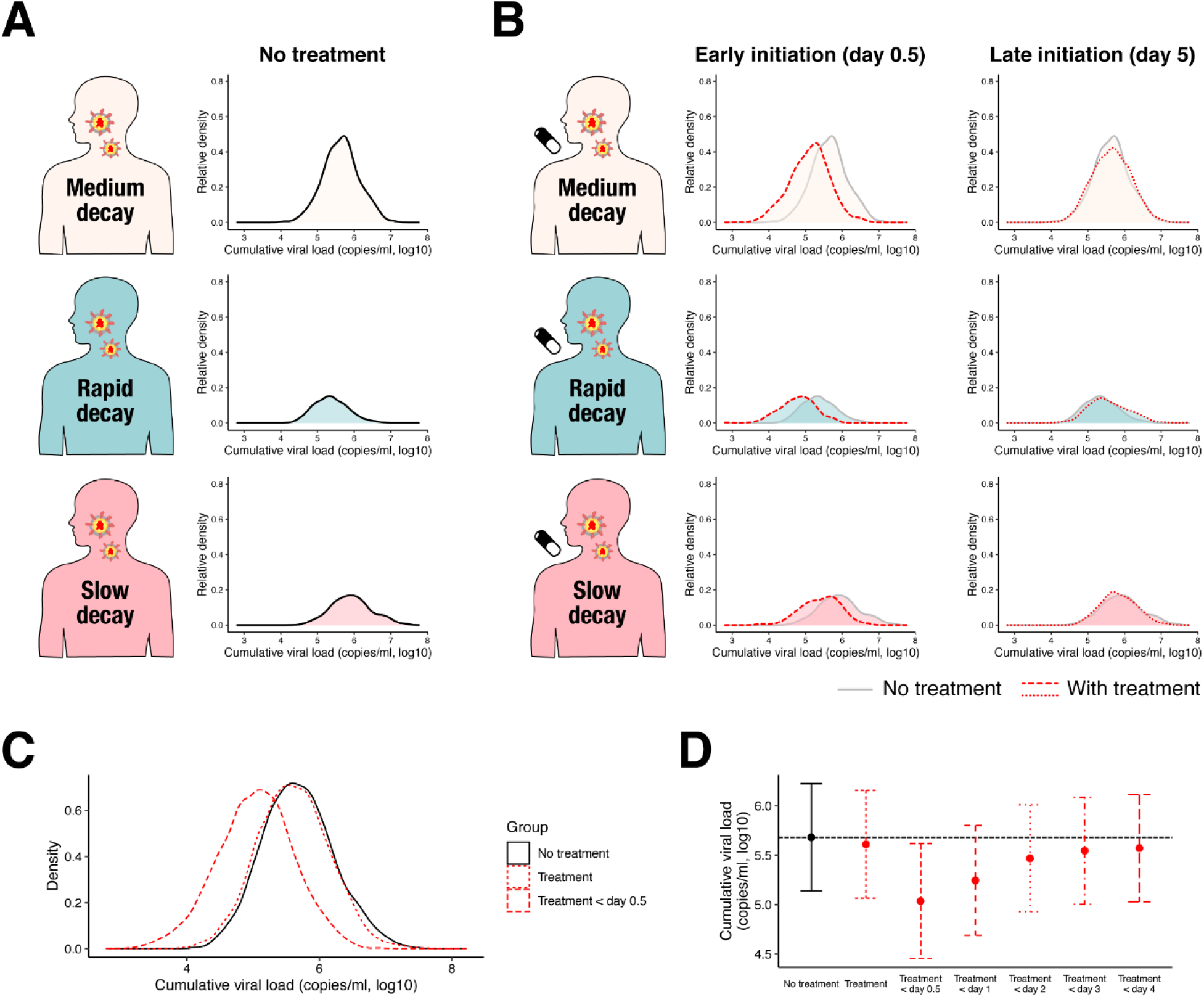
Cumulative viral load in different types of patients. Distributions of cumulative viral load (in log scale) in each patient group without and with the treatment (the treatment is initiated at 0.5 [‘Early’] or 5 [‘Late’] days after onset of symptoms) were shown in (A) and (B), respectively. The distributions are represented as ‘relative density’ to reflect different proportions of the three groups. (C) The distributions of cumulative viral load for patients without and with treatment were shown in black and red curves, respectively. The two treatment strategies were implemented: (Treatment) patients were treated immediately after hospitalization regardless of the time from onset to enrollment, (Treatment < day 0.5) patients were treated immediately after hospitalization if they are hospitalized within 0.5 days from the onset of symptoms. (D) Means and standard deviation of cumulative viral load under different inclusion criteria are shown.

**Table S1.** Summary of clinical trials for antivirals for SARS-CoV-2.

| Treatment | Study design | Timing of initiation (days) | Sample size (control/treatment) | Primary outcome* | Effective<sup>#</sup> | Reference | Peer-reviewed |
| --- | --- | --- | --- | --- | --- | --- | --- |
| Lopinavir/ritonavir | RCT | 13 (IQR: 11-16) | 99/100 | 1 | No | (13) | Yes |
| Remdesivir | RCT | 10 (IQR: 9-12) | 79/158 | 1 | No | (14) | Yes |
| Hydroxychloroquine | RCT | - | 31/31 | 5 | Yes | (15) | No |
| Hydroxychloroquine | RCT | - | 75/75 | 3 | No | (16) | Yes |
| Favipiravir | RCT | - | 120/120 | 1 | No | (17) | No |
| Lopinavir/ritonavir | RCT | 3.3 (SD: 1-15) | 21/7 | 3 | No | (18) | No |
| Arbidol | RCT | 3.2 (SD: 0.5-11) | 16/7 | 3 | No | (18) | No |
| Favipiravir | OS | <7 | 45/35 | 3 | Yes | (19) | Yes |
| Remdesivir | OS | 12 (IQR: 9-15) | 53 | 2 | - | (20) | Yes |
| Hydroxychloroquine | OS | 4 (SD: 2-6) | 16/20 | 3 | Yes | (21) | Yes |
| Hydroxychloroquine | OS | 4.9 (SD: 1.3-8.5) | 80 | 4 | - | (4) | Yes |
| Meplazumab | OS | - | 17/10 | 3 | Yes | (22) | No |

\* 1. Clinical improvement/recovery, 2. Oxygen-support class, 3. Virological clearance, 4. (i) Clinical course, (ii) Contagiousness, (iii) Length of stay in the infectious disease ward, 5. Time to clinical recovery.
<sup>#</sup> Whether the primary outcome is statistically significantly different between control and treatment group. ‘-’ denotes no statistical test was performed because it is a single arm study (i.e., no control group).

**Table S2.**
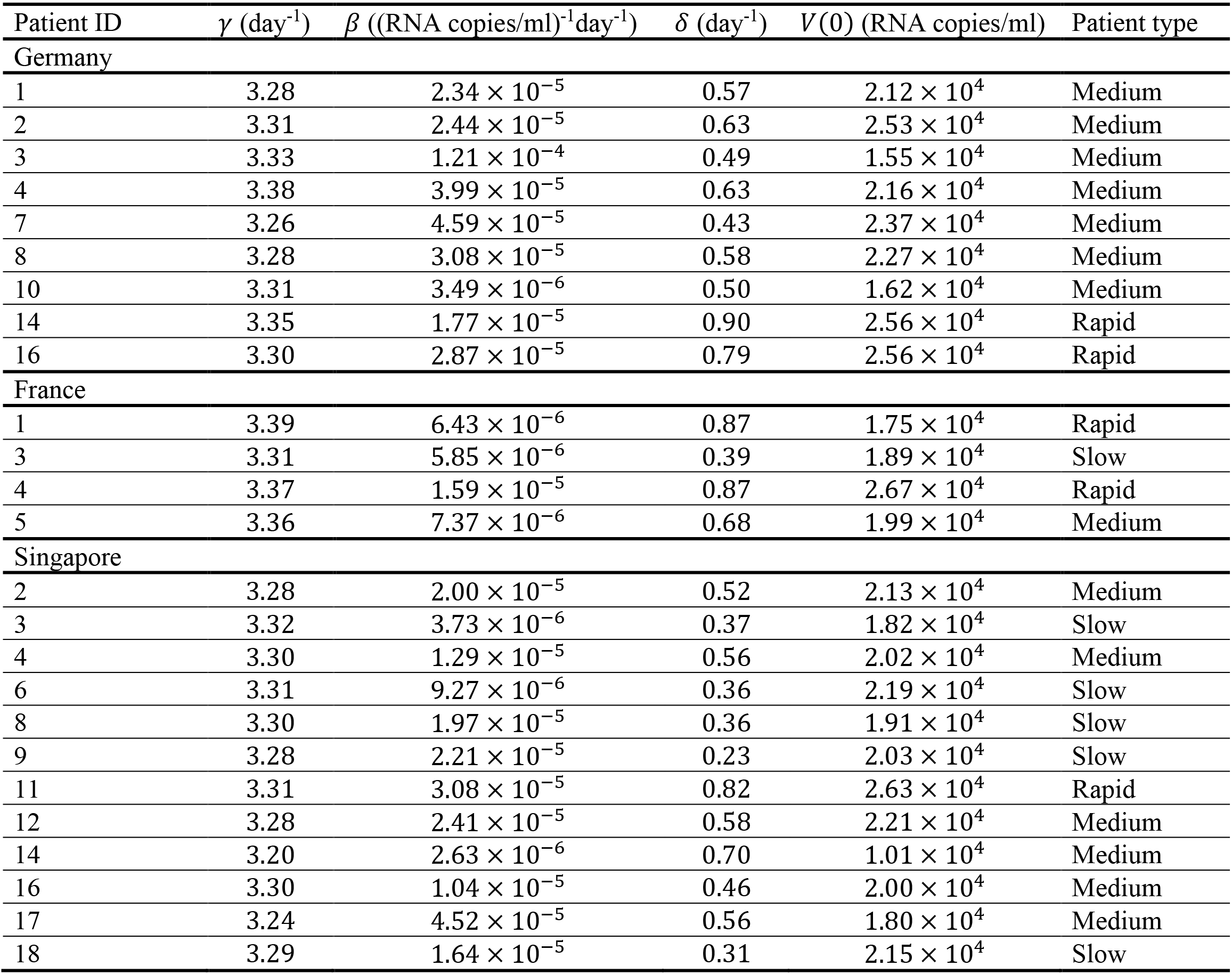
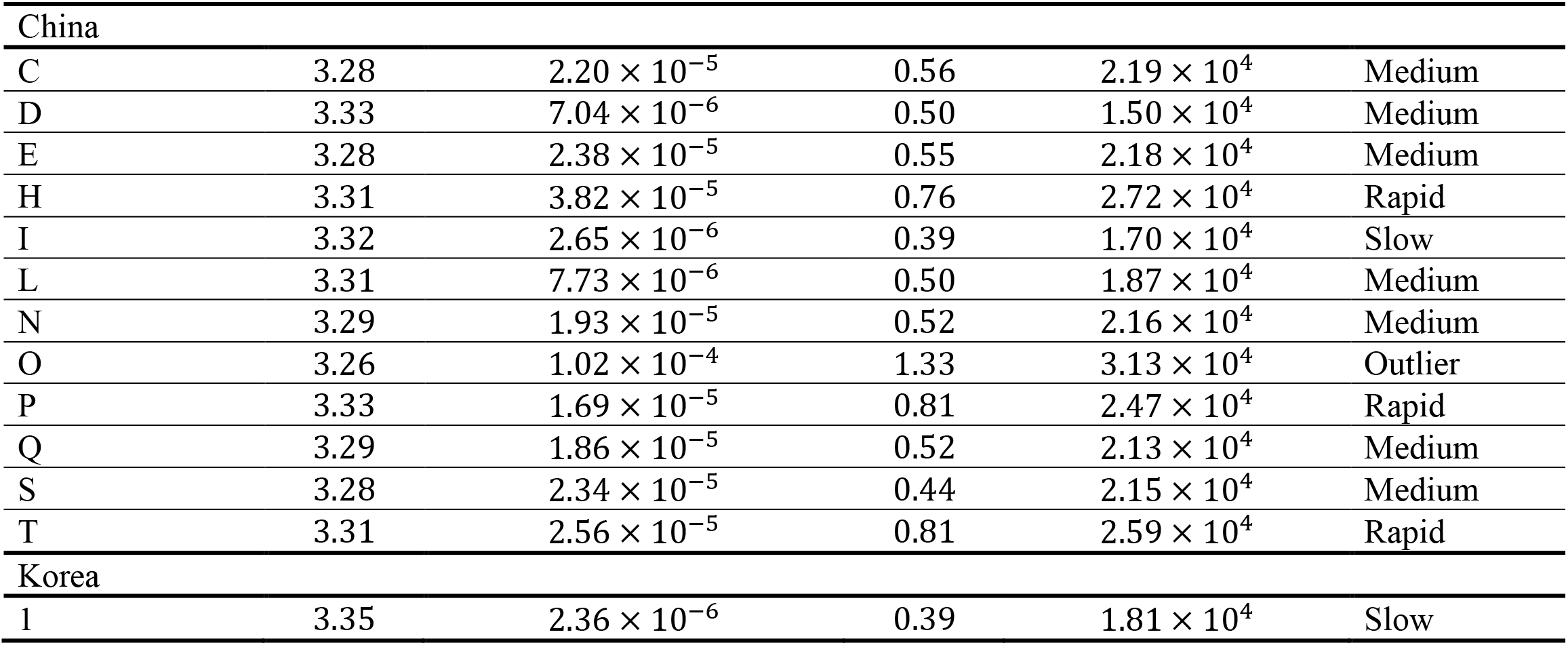
Estimated parameters for each patient.

| Patient ID | $\gamma$ (day<sup>-1</sup>) | $\beta$ ((RNA copies/ml)<sup>-1</sup>day<sup>-1</sup>) | $\delta$ (day<sup>-1</sup>) | $V(0)$ (RNA copies/ml) | Patient type |
| --- | --- | --- | --- | --- | --- |
| Germany |  |  |  |  |  |
| 1 | 3.28 | $2.34 \times 10^{-5}$ | 0.57 | $2.12 \times 10^4$ | Medium |
| 2 | 3.31 | $2.44 \times 10^{-5}$ | 0.63 | $2.53 \times 10^4$ | Medium |
| 3 | 3.33 | $1.21 \times 10^{-4}$ | 0.49 | $1.55 \times 10^4$ | Medium |
| 4 | 3.38 | $3.99 \times 10^{-5}$ | 0.63 | $2.16 \times 10^4$ | Medium |
| 7 | 3.26 | $4.59 \times 10^{-5}$ | 0.43 | $2.37 \times 10^4$ | Medium |
| 8 | 3.28 | $3.08 \times 10^{-5}$ | 0.58 | $2.27 \times 10^4$ | Medium |
| 10 | 3.31 | $3.49 \times 10^{-6}$ | 0.50 | $1.62 \times 10^4$ | Medium |
| 14 | 3.35 | $1.77 \times 10^{-5}$ | 0.90 | $2.56 \times 10^4$ | Rapid |
| 16 | 3.30 | $2.87 \times 10^{-5}$ | 0.79 | $2.56 \times 10^4$ | Rapid |
| France |  |  |  |  |  |
| 1 | 3.39 | $6.43 \times 10^{-6}$ | 0.87 | $1.75 \times 10^4$ | Rapid |
| 3 | 3.31 | $5.85 \times 10^{-6}$ | 0.39 | $1.89 \times 10^4$ | Slow |
| 4 | 3.37 | $1.59 \times 10^{-5}$ | 0.87 | $2.67 \times 10^4$ | Rapid |
| 5 | 3.36 | $7.37 \times 10^{-6}$ | 0.68 | $1.99 \times 10^4$ | Medium |
| Singapore |  |  |  |  |  |
| 2 | 3.28 | $2.00 \times 10^{-5}$ | 0.52 | $2.13 \times 10^4$ | Medium |
| 3 | 3.32 | $3.73 \times 10^{-6}$ | 0.37 | $1.82 \times 10^4$ | Slow |
| 4 | 3.30 | $1.29 \times 10^{-5}$ | 0.56 | $2.02 \times 10^4$ | Medium |
| 6 | 3.31 | $9.27 \times 10^{-6}$ | 0.36 | $2.19 \times 10^4$ | Slow |
| 8 | 3.30 | $1.97 \times 10^{-5}$ | 0.36 | $1.91 \times 10^4$ | Slow |
| 9 | 3.28 | $2.21 \times 10^{-5}$ | 0.23 | $2.03 \times 10^4$ | Slow |
| 11 | 3.31 | $3.08 \times 10^{-5}$ | 0.82 | $2.63 \times 10^4$ | Rapid |
| 12 | 3.28 | $2.41 \times 10^{-5}$ | 0.58 | $2.21 \times 10^4$ | Medium |
| 14 | 3.20 | $2.63 \times 10^{-6}$ | 0.70 | $1.01 \times 10^4$ | Medium |
| 16 | 3.30 | $1.04 \times 10^{-5}$ | 0.46 | $2.00 \times 10^4$ | Medium |
| 17 | 3.24 | $4.52 \times 10^{-5}$ | 0.56 | $1.80 \times 10^4$ | Medium |
| 18 | 3.29 | $1.64 \times 10^{-5}$ | 0.31 | $2.15 \times 10^4$ | Slow |
| China |  |  |  |  |  |
| C | 3.28 | $2.20 \times 10^{-5}$ | 0.56 | $2.19 \times 10^4$ | Medium |
| D | 3.33 | $7.04 \times 10^{-6}$ | 0.50 | $1.50 \times 10^4$ | Medium |
| E | 3.28 | $2.38 \times 10^{-5}$ | 0.55 | $2.18 \times 10^4$ | Medium |
| H | 3.31 | $3.82 \times 10^{-5}$ | 0.76 | $2.72 \times 10^4$ | Rapid |
| I | 3.32 | $2.65 \times 10^{-6}$ | 0.39 | $1.70 \times 10^4$ | Slow |
| L | 3.31 | $7.73 \times 10^{-6}$ | 0.50 | $1.87 \times 10^4$ | Medium |
| N | 3.29 | $1.93 \times 10^{-5}$ | 0.52 | $2.16 \times 10^4$ | Medium |
| O | 3.26 | $1.02 \times 10^{-4}$ | 1.33 | $3.13 \times 10^4$ | Outlier |
| P | 3.33 | $1.69 \times 10^{-5}$ | 0.81 | $2.47 \times 10^4$ | Rapid |
| Q | 3.29 | $1.86 \times 10^{-5}$ | 0.52 | $2.13 \times 10^4$ | Medium |
| S | 3.28 | $2.34 \times 10^{-5}$ | 0.44 | $2.15 \times 10^4$ | Medium |
| T | 3.31 | $2.56 \times 10^{-5}$ | 0.81 | $2.59 \times 10^4$ | Rapid |
| Korea |  |  |  |  |  |
| 1 | 3.35 | $2.36 \times 10^{-6}$ | 0.39 | $1.81 \times 10^4$ | Slow |

**Table S3.** Sample size (per group) under different inclusion criteria.

| Outcome | All patients<br>(no inclusion criteria) | Patients treated in “X days” from the onset of symptoms |  |  |  |  |
| --- | --- | --- | --- | --- | --- | --- |
|  |  | 0.5 days | 1 days | 2 days | 3 days | 4 days |
| Duration of virus shedding | 2720 | 98 | 204 | 866 | 1054 | 2085 |
| Cumulative viral load | 1862 | 26 | 52 | 206 | 503 | 774 |

